# Primary Care Providers’ Perspectives on Receiving Tier 1 Genomic Results from a National Study – the Million Veteran Program Return Of Actionable Results (MVP-ROAR) Study

**DOI:** 10.1101/2024.10.29.24316065

**Authors:** Anna L. Johannsen, Morgan E. Danowski, Kailyn E. Sitter, Charlene L. Preys, Haley C. Gerety, Charles A. Brunette, Kurt D. Christensen, J. Michael Gaziano, Joshua W. Knowles, Sumitra Muralidhar, Amy C. Sturm, Yan V. Sun, Stacey B. Whitbourne, Thomas Yi, the VA Million Veteran Program, Jason L. Vassy

**Affiliations:** VA Boston Healthcare System, Boston, MA, USA; Harvard Medical School, Department of Medicine, Boston, MA, USA; PRecisiOn Medicine Translational Research Center, Department of Population Medicine, Harvard Pilgrim Health Care Institute; Stanford University, School of Medicine, Palo Alto, CA, USA; Department of Veterans Affairs, Office of Research and Development, Washington D.C.; 23andMe Inc., Sunnyvale, CA; Atlanta VA Healthcare System, Decatur, GA, USA; Department of Epidemiology and Global Health, Emory University Rollins School of Public Health, Atlanta, GA, USA

## Abstract

**Background:** Patients are increasingly obtaining genetic health information and integrating it into their care with the help of their primary care provider (PCP). However, PCPs may not be adequately prepared to effectively utilize genetic results . Across the VA health system, the Million Veteran Program-Return of Actionable Results-Familial Hypercholesterolemia (MVP-ROAR) study clinically confirms and returns genetic results associated with familial hypercholesterolemia (FH), identified in a national biobank program.

**Methods:** PCPs who received their patient’s genetic results through the MVP-ROAR Study were invited to participate in semi-structured interviews, which explored PCPs’ familiarity with FH, how the results impacted medical management, and suggestions for process improvement. Interviews were transcribed and analyzed using directed content analysis and constant comparison methods to identify key themes.

**Results:** Interviews with nine PCPs revealed varied levels of familiarity with genetic testing and FH. Most PCPs did not distinguish FH from common high cholesterol issues and already used similar treatment approaches. Many PCPs did not recall receiving results from the MVP-ROAR Study. Alerts in medical records were deemed effective for communicating results. PCPs valued genetics in informing patient care and identifying at-risk family members but noted several implementation barriers, such as additional workload and unclear medical management benefits. Recommendations for improving results disclosure included simplifying the genetic testing report and associated support documents.

**Conclusion:** The study represents the first investigation into PCPs’ experiences with receiving genetic test results from a biobank linked to a national healthcare system. Results suggest that PCPs generally view genetic testing as beneficial, though they may not significantly alter medical management. PCPs expressed that integrating genetics into routine care may be burdensome and require additional training, which may not be practical. The study underscores the need for accessible genetic information, which could be aided by specialized support roles or different clinical specialties assisting with incorporating genetic results into patient care.

## INTRODUCTION

Individuals increasingly have opportunities to receive health-related genomic results, including through direct-to-consumer or research offerings, cascade screening recommended as a result of their relatives’ genetic results, or secondarily from genetic testing ordered for other clinical indications.^1–3^ At the same time, there is growing interest in population-wide genomic screening for certain conditions that are sufficiently prevalent, highly penetrant, and clinically actionable to justify their widespread detection and management.^4,5^ Indeed, recent modeling studies suggest that population screening for three Centers for Disease Control and Prevention (CDC) Tier 1 genomic conditions—hereditary breast and ovarian cancer, Lynch syndrome, and familial hypercholesterolemia—may be cost-effective at the societal level.^6,7^

As genomic testing moves increasingly from specialty care into population screening, primary care providers (PCPs) will find themselves on the front line of this new clinical paradigm. Studies have reported that PCPs feel unprepared for and unsupported in helping their patients interpret and manage genomic results, especially if they were not the ordering provider.^8,9^ Nevertheless, PCPs value their primary role in managing and coordinating their patients’ care and desire systems that support the appropriate interpretation and management of genomic results.^10,11^ On one hand, they appreciate delegating certain clinical tasks to other staff members or technological platforms to streamline procedures;^12,13^ on the other hand, they wish to remain sufficiently informed to effectively direct their patients’ care.^14^

In this context, it is crucial to understand PCPs’ perspectives on receiving unanticipated yet actionable genomic results for their patients and on the processes that can support them in managing these results appropriately. Despite expressing limited bandwidth to integrate genetic research results into clinical practice, PCPs remain central in managing multiple facets of their patients’ care. To better inform the future integration of genomic testing and screening into primary care, we conducted an interview study among a national sample of PCPs who received unexpected results for their patients participating in a mega-biobank and national return-of-results pilot project.

## METHODS

### MVP-ROAR Study

This interview study is a substudy of the Million Veteran Program Return Of Actionable Results (MVP-ROAR) Study, described previously.^16^ In brief, the Million Veteran Program (MVP) is a national biobank that to date has enrolled more than 1 million US military veterans, who complete surveys, provide DNA specimens, and consent to the research use of their medical record data.^15^ Within MVP, MVP-ROAR is a pilot trial among participants whose MVP research genotype data suggest they carry a pathogenic variant associated with familial hypercholesterolemia (FH). Familial hypercholesterolemia was selected as the exemplar genetic condition for MVP-ROAR because it is recognized by the American College of Medical Genetics and Genomics as a reportable secondary finding and by the CDC as a Tier 1 condition and because it has associated cholesterol-lowering treatment guidelines easily implementable by PCPs.^16^ In the MVP-ROAR Study, MVP participants with a potential FH-associated variant were recontacted by the MVP-ROAR genetic counselor and invited to undergo confirmatory clinical gene sequencing and post-test genetic counseling, including provision of FH-related informational resources and facilitation of cascade testing for at-risk family member. Each participant’s PCP was also sent the results via email, along with an FH treatment algorithm^16^ and a letter summarizing their patient’s results.^17^ No action was required from PCPs by the study, but they were encouraged to discuss the result and management recommendations with their patients. The study genetic counselor was available as an ongoing resource to both participants and PCPs.

### PCP interview study

To capture PCPs’ perceptions, preferences, and needs regarding the receipt of clinical genomic results, we conducted a qualitative interview study among a national sample of PCPs who had received clinical gene panel sequencing results from the MVP-ROAR Study for at least one of their patients. The analytic team included one GC student (AJ), three GCs (MD, CP, HL), and one medical anthropologist (KS). The VA Central Institutional Review Board approved this substudy to the MVP-ROAR protocol (#19-11).

#### Participants and recruitment

PCPs were eligible to participate in the interview substudy if 1) they practiced primary care at a Veterans Health Administration location, 2) their patient received genetic test results through the MVP-ROAR Study, and 3) the patient had completed the end-of-study survey. To recruit our sample, study staff (AJ, MD) invited eligible PCPs (n=52) to participate using emails and internal institutional instant messages.

#### Interview design and data collection

We set out to understand 1) the impact of receiving these genomic results on patient care; 2) PCPs’ comfort and preparedness in discussing these results with their patients; 3) perceived facilitators and barriers to receiving results within a clinical research framework; and 4) suggestions for improving the results communication process. Our interview guide was created through an iterative process using *a priori* constructs from the Consolidated Framework for Implementation Research (CFIR),^18^ which offers researchers a basis for understanding potential barriers and facilitators to successful implementation^19^ of new initiatives. A two-person team (AJ, MD) conducted semi-structured interviews using video software, Microsoft Teams, from August 2023 to February 2024. Researchers took detailed written notes during the interviews, and all interviews but one were audio recorded and transcribed verbatim. In the sole case of PCP 3, where the interview was not audio recorded, researchers used their notes in place of a transcript. Because PCP receipt of original genomic results occurred 2-34 months prior to this substudy, each PCP was re-sent their patient’s genetic testing results report, the summary letter explaining the implications of the patient’s results, and the FH treatment algorithm before their interview. The interviews asked PCPs about their 1) familiarity with FH; 2) experience receiving results through the MVP-ROAR Study; 3) discussion of results with patients; 4) recommendations for improving the MVP-ROAR Study results communication process; 5) general impressions of genetic testing; and 6) facilitators and barriers of genetic testing both within the MVP-ROAR Study and more broadly. We also collected information about demographics, including whether PCPs had received any formal or informal genetics training. Due to the narrow focus of the interviews, our team reached thematic saturation with a smaller sample size than is typically required.^20,21^ Interviews lasted approximately 30 minutes.

#### Data analysis

We conducted a directed content analysis ^22,23^ to understand how PCPs experience the receipt of results and implement and/or discuss these results with their patients who have participated in the MVP-ROAR program. Using a rapid qualitative analysis approach, coders (AJ, MD) reviewed the interview notes and sorted interview data into a matrix based on *a priori* categories^18^ created using CFIR and feedback from co-authors. One researcher (KS) advised on the analytical approach. Two additional researchers (HL, CP) assisted the coders with creating iterative summaries of the categorized data across interviews. Summaries were reviewed for consistency and differentiation.^24^ The full team met regularly to discuss data, refine the approach, and add emergent categories drawn from the data about PCPs’ experiences and the barriers and facilitators they encountered when engaging with the MVP-ROAR program, as not all data collected fit within the *a priori* categories.^25,26^ We also utilized constant comparison methods when reviewing data summaries to identify and understand the breadth of PCP experiences as they received genomic test results from the MVP-ROAR program. Our team reached consensus on the final data categorization, data summaries, and interpretation through weekly analysis meetings. Engaging all members of the core research team (AJ, MD, CP, HL, KS) in these meetings also reduced bias in data interpretation.

## RESULTS

We interviewed 9 PCPs (Table 1) from VA facilities across 7 states (Figure 1). The following themes emerged from these interviews.

**Table 1.** Characteristics of 9 primary care providers interviewed about their experiences receiving genomic results from the Million Veteran Program Return Of Actionable Results Study.

| **Characteristic** | ***n* (%)** |
| --- | --- |
| Gender |  |
| Male | 3 (33) |
| Female | 5 (56) |
| Not provided | 1 (11) |
| Race |  |
| White | 3 (33) |
| Black or African American | 1 (11) |
| Asian | 4 (44) |
| Not provided | 1 (11) |
| Years in primary care |  |
| < 10 years | 3 (33) |
| 10 – 14 years | 0 (0) |
| 15 – 19 years | 1 (11) |
| 20 – 25 years | 2 (22) |
| > 25 years | 3 (33) |
| Years working for VA |  |
| < 10 years | 5 (56) |
| 10 – 19 years | 2 (22) |
| 20 – 30 years | 1 (11) |
| > 30 years | 1 (11) |
| Region of VA facility |  |
| Southeast | 4 (44) |
| Northeast | 2 (22) |
| Midwest | 1 (11) |
| West | 2 (22) |
| Genetics training beyond medical school |  |
| None | 7 (78) |
| Reported some | 2 (22) |
| Individual patient results received |  |
| 1 | 8 (89) |
| 2 | 1 (11) |

**Figure 1.**
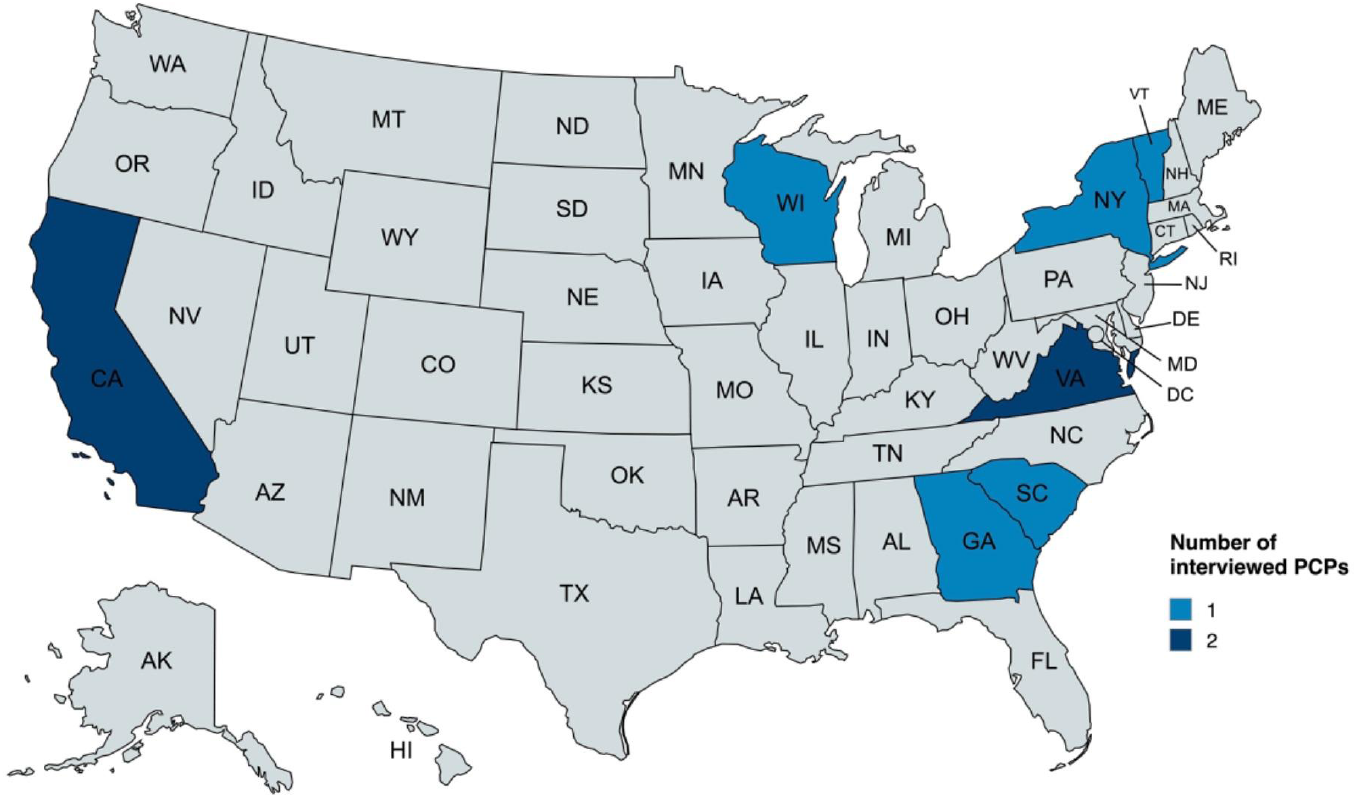
VA locations of 9 primary care providers interviewed about their experiences receiving genomic results from the Million Veteran Program Return Of Actionable Results Study.

### Familiarity with genetic testing and FH

Most reported having no previous formal or informal genetics training; some discussed attending occasional genetics educational talks hosted by the VA, often related to the VA National Pharmacogenomics Program (formerly the PHASER program).^27^ PCP familiarity with FH varied. While some were familiar with FH given their own family or patients’ histories, many did not distinguish it from multifactorial high cholesterol: “…I don’t actively differentiate in terms of if [high cholesterol is] familial, we just diagnose and treat hyperlipidemia…” [PCP 8]. Regardless, PCPs expressed confidence in treating high cholesterol and hyperlipidemia, some describing these conditions their “bread and butter” [PCPs 1 and 6].

### Experience receiving results

Most PCPs did not recall receiving their patients’ MVP-ROAR Study results via email, although they reported that this would generally be an effective way to receive such results. PCPs noted that the result report was too lengthy, contained confusing medical jargon, and “difficult to read.” If they did remember receiving the results package, PCPs reported it did not impact their management of the patient’s clinical care, for which they reported they already use similar treatment algorithms. One provider, with 21 years of experience, felt the results and accompanying resources might be helpful to early-career PCPs or to those less familiar with the difference between multifactorial high cholesterol and FH; another provider felt the information about clinical and family implications seemed particularly useful due to implications for cascade testing. One provider described the treatment algorithm as helpful, and planned to use it when treating other patients.

### Impact of MVP-ROAR results

The PCPs reported that the FH results did not impact their care of MVP-ROAR participants. Many reported already effectively managing their patients’ cholesterol levels prior to receiving results, including having discussions with patients about medication changes, dietary recommendations, family histories, and referrals to specialists. One PCP asked, “What is the benefit of doing the testing if I’m not changing any management?” [PCP 9]. PCPs generally agreed that receiving genetic testing results could theoretically inform medical management and treatment of patients, identify at-risk family members, or provide access to novel, targeted treatments, but recognized the potential limitations of prescribing within the VA:

> “The difficulty, at least in the primary care sector, is just fighting all the noise about medications and treatment of [FH or other lipid disorders]…you start getting into treatment and that’s where a lot of resistance comes into play, because [patients] hear about statins, which are typically the first line, and here at the VA, PCSK9 inhibitors require authorization.” [PCP 2]

Some PCPs, unsurprised by their patients’ genetic results, expressed gratitude for finally having an explanation for why their patients’ cholesterol levels were challenging to treat and responded poorly to statins. However, FH test results were unexpected but welcome for a PCP who gained new perspective about one of their patients: “I actually think it’s great… for this particular patient, I was definitely not aware of this diagnosis [… so this] helps me to focus a little bit more on another problem she has that I was less aware of” [PCP 5].

Management of FH may involve referral to specialists if elevated cholesterol levels remain or are especially treatment resistant. One provider described referring “patients who need more [advanced management] to cardiology,” although they mentioned knowing “what to do with statins” and managing most of their hyperlipidemia patients in primary care [PCP 4]. When caring for patients who did not respond sufficiently to usual statins, PCPs reported they most commonly referred to cardiology and lipid specialists, although some PCPs also referred patients to endocrinologists and clinical pharmacists.

### Expanding return of results

PCPs saw theoretical value in receiving genetic testing results for FH and stated interest in seeing genetic results disclosure expanded to other conditions, especially hereditary cancers, diabetes, and hypertension. One provider shared, “I think this is where medicine is headed … I think this genetic pathway is something that we should be pursuing a lot more aggressively” [PCP 2]. Another provider, who sees only female-identifying patients, said their patients would “welcome that extra eye on them, especially for family history or [if] they’re stressed out about [the hereditary components of breast cancer]” [PCP 1]. Other benefits of expanded return of results programs include providing evidence to support PCPs’ clinical recommendations, encouraging patients to remain compliant in their screenings or treatments, and motivating patients to make positive lifestyle changes. Additionally, negative results may alleviate patients’ concerns about developing a familial condition: “If I can give them a negative result or something not to worry about, that really helps with the mental health, and [PCPs] spend a lot of time dealing with the mental health aspect.” [PCP 1].

Discussions of how valuable expanded genetic results disclosure could theoretically be paired with discussions of barriers to implementing expansion of a program like MVP-ROAR. PCPs were primarily concerned with adding more time-consuming work into their already hectic workloads because they are “…so, so busy, so overwhelmed” [PCP 6]. This feeling was exacerbated by concerns about whether genetic test results might “[translate] to better clinical care” while “add[ing] more work to providers” [PCP 8] since they were uncertain how test results might directly impact medical management of patients for most conditions. PCPs largely felt they did not have expertise to identify appropriate testing candidates, order testing, or interpret and disclose results. In addition, time constraints during appointments limit PCPs’ ability to include conversations about genetic testing:

> “[In] primary care we have 20, 25 different complaints to take care of with limited time, like 30 minutes. So, we don’t get to explore more…sometimes we [schedule their next appointment] sooner and have that discussion; sometimes, we end up calling them after hours and talk about it.” [PCP 6]

Providers described mixed patient views of genetic testing, from interested to ambivalent to hesitant. They generally agreed that most patients who may have reason to consider genetic testing (*i*.*e*., personal or family history of disease) seem interested or at least open to pursuing genetic testing, particularly surrounding cancer risks. PCP 1 described, “…[T]hey welcome that extra eye on them, especially for family history or [if] they’re stressed out about it.” Alternately, PCPs reported some patients’ negative views of genetic testing and concerns about privacy, discrimination, or undue stress, fear, and uncertainty about potential future symptoms as additional barriers to expanding test results disclosure. One provider described, “It could cause great stress knowing that you have a genetic disorder. Some people don’t want to know if they’ll get a disease, others want to know and try to prevent it, or look for it, so every person’s different,” [PCP 7]. One PCP cautioned that genetic testing should only be done on individuals who are presumed to be affected, because general population screening may cause unnecessary fear; another thought expansion should only include later-onset conditions where some level of prevention can be implemented. Additionally, three PCPs cited financial concerns as reasons patients outside the VA may not be receptive to genetic testing: “…We would have never entertained technology like this in private practice because [of the] cost and insurance companies wouldn’t pay for it” [PCP 1]. PCPs were concerned that insurance coverage (or lack thereof) may leave patients with a hefty bill or limited access to genetic testing or necessary follow-up procedures.

### Recommendations for improvement

PCPs disagreed about whether additional resources included in the results disclosure package might be helpful to them. Some mentioned a desire for additional education and support in managing FH patients, such as continuing medical education sessions provided by the genetics department, while others were unsure if they would utilize resources due to time constraints. PCPs wanted future return of results packages to include minimal jargon, be in an easy-to-read layout with the most important information first in a clearly labeled main section, and make clear what information is supplemental and not critical for medical management. They stated the organization of the documents was crucial, so that PCPs would not spend time reading extraneous information or miss more important information located elsewhere in the results document. Alerts in the medical record, which are prominently displayed within the software, were identified as the most effective way to communicate genetic test results to PCPs. Alerts are available to each of the patient’s providers, even if the patient receives care from providers at multiple VA locations. PCPs generally agreed that e-mail, VA internal instant messaging, and mail could be used to communicate test results if necessary

## DISCUSSION

This study examined PCPs’ actual experiences receiving their patients’ genomic results for a CDC Tier 1 condition from a biobank linked to a national integrated healthcare system. PCPs generally reported clinical benefit in receiving positive FH-associated variant results. On the other hand, they paradoxically did not find that the results necessitated changes in their patients’ medical management, particularly in cases where they reported they had already been effectively managing their patients’ hypercholesterolemia. PCPs also highlighted the time-constrained nature of their primary care practice as a barrier to integrating genetics into routine clinical care. They recommended that future genomic screening initiatives might be enhanced through the communication of results and easy-to-understand supportive information via common clinical communication methods like internal email and instant messaging and EHR alerts. They suggested that additional genetics training or continuing education might be beneficial but also infeasible with their demanding clinical schedules. With supports in place, PCPs perceived value in receiving genomic results from an expanded list of conditions beyond FH.

Previous studies have examined PCPs’ perspectives on receiving unsolicited genomic results, but these have generally been hypothetical in nature.^28–31^ For example, in an interview study at four healthcare systems preparing to participate in large-scale return-of-results project, physicians responded in the abstract about the need for actionability, evidence-based management plans, and clinical decision support when receiving unsolicited genomic results for their patients.^29^ At another healthcare system preparing to return genomic results to biobank participants, a survey of PCPs indicated they had a desire to receive results, preferably by EHR or letter, but that the majority wanted a genetics specialist to be involved in communicating results to patients. Still, about half reported that the PCP should share the responsibility of discussing the results.^30^ The present study extends this work by interviewing a national sample of PCPs after they had received real genomic results for at least one of the patients in their primary care panel. Perhaps because of this real experience, PCPs in this study did not express theoretical concerns about the program but instead focused more on how to improve the implementation of such a program into busy primary care practice.

These findings thus have practical implications for future population screening programs for FH specifically and other genomic conditions more generally. The majority of patients who receive primary care at a VA facility are middle aged or older and thus have multiple medical conditions, including high cholesterol and other cardiovascular disease risk factors.^32,33^ Lipid panel testing is ordered frequently in the VA, and many, but not all, VA patients with hypercholesterolemia or elevated cardiovascular disease risk receive adequate therapy and achieve target LDL cholesterol levels.^34–36^ This may explain the perception among PCPs interviewed in this study that an FH-associated genomic result would not significantly change their clinical management of carriers. Evidence from the VA and other healthcare systems, however, suggests that patients with an FH phenotype may go undetected and undertreated ^35,37,38^ for their level of cardiovascular disease risk. To realize the benefits of intensive lipid-lowering for patients with a molecular diagnosis of FH, let alone the benefits of cascade testing among their relatives,^39,40^ genomic screening programs will need to communicate the importance of distinguishing between FH and more common forms hypercholesterolemia and support PCPs in making clinical decisions based on those distinctions.

When asked about the expansion of genomic screening to additional conditions, PCPs in the present study indicated that screening for hereditary cancer risk would be beneficial. This preference may stem from the comparative difficulty of assessing cancer risk, which unlike hypercholesterolemia with a readily measurable biomarker, relies on data such as detailed family histories that are more challenging to collect and interpret comprehensively in the time-constrained environment of primary care practice. Although not asked specifically, this finding might indicate acceptance of screening for hereditary breast and ovarian cancer and Lynch syndrome, two other Tier 1 conditions. Nonetheless, PCPs in this and other studies consistently report feeling inadequately prepared and lacking sufficient time to independently receive, interpret, and manage genomic screening results.^31,41^ To address these challenges, novel service delivery models, such as delegation of some tasks to ancillary staff, traditional or electronic consult services, or other population management strategies, have been implemented in various healthcare systems to facilitate genomic medicine care.^12,13^ Digital tools might also play a role in supporting both the patient and PCP in appropriate management.^42–45^ A combination of these and other approaches will likely be essential for the meaningful integration of genomic screening into primary care. Healthcare systems should include frontline PCPs in the codesign of workflows and support systems for future genomic screening initiatives.

This study has strengths and limitations. It reports the actual experiences among a national sample of PCPs receiving real genomic screening results. The views of the PCP participants are not necessarily generalizable to all PCPs; participants indicated willingness to be interviewed about their experiences with the MVP-ROAR Study, which may indicate that they were more willing to integrate genomic screening into their care or that they had strong opinions about the program. However, the variation among participants’ responses suggests that we captured a range of opinions and experiences, a goal of qualitative research seeking to inform implementation.^46^ There was a prolonged period of time between some PCPs’ receipt of their patients’ results and their interviews, which may have impacted their ability to recall their initial reactions to receiving the results. To minimize this impact, results and study informational resources were provided again to each PCP to review in preparation for their interview.

## Conclusion

This study underscores the practical need for enhanced support and infrastructure in genomic screening within primary care. If PCPs are to navigate the complexities of integrating genomic results into patient management, clear communication and support systems including collaborative care approaches are essential for the effective use of genomic data in improving patient outcomes. Moving forward, healthcare systems must prioritize the development of integrated strategies that address PCPs’ concerns and workload, facilitating the broader application of genomic screening in routine clinical practice.

## Supporting information

MVP-ROAR PCP Interview Guide

MVP Core Acknowledgements

## Data Availability

All data produced in the present study are available upon reasonable request to the authors.

## Acknowledgements

This research is based on data from the Million Veteran Program, Office of Research and Development, Veterans Health Administration, and was supported by award MVP030. This publication does not represent the views of the Department of Veteran Affairs or the United States Government. The authors thank the following for their assistance with the manuscript: Ella Ransbottom, Annika Toivonen, Camille Amaditz, and Rosalina Caliri.

