## Supplementary material for "Primary Care Providers’ Perspectives on Receiving Tier 1 Genomic Results from a National Study – the Million Veteran Program Return Of Actionable Results (MVP-ROAR) Study": MVP-ROAR PCP Interview Guide

[INTERVIEWER INTRODUCTION] *include role at VHA and purpose of interview*.

[START RECORDING]

You have been invited to participate in a research interview about the Million Veteran Program - Return of Actionable Results (MVP-ROAR) Study. One or more of your patients enrolled in MVP-ROAR Study, the first return-of-results project in MVP. Because of your patient's participation in the study, you received their genetic test results from the research team. We are doing this interview study to learn more about your experience receiving those results.

Your participation is voluntary. Your responses will remain confidential and will not be shared with your supervisor. You may stop the interview at any time, and you do not have to answer any question you do not wish to answer. We will record and transcribe this interview. All recordings and transcripts will be stored in an encrypted database accessible only to authorized researchers.

Do you have any questions for me?

Before we begin, please state your name and today's date. [PCP states name and date]

Do you consent to today's interview? [PCP states yes or no for the recording. If no, the interview will be terminated]

Do I have your permission to record the interview? [PCP states yes or no for the recording]

Thank you. First, I'd like to ask you a few questions about your experience as a VA primary care provider.

Professional role

1. How long have you been practicing primary care?
2. How long have you been practicing at the VA? Which station(s)?
3. Have you had any formal/informal genetics training? If you have, can you please describe these training experiences?

MVP-ROAR [INTERVIEWER USES THESE PROMPTS TO EXPLORE PCP EXPERIENCE WITH MVP-ROAR BUT MAY ADAPT THE PROMPTS BASED ON THE SPECIFIC EXPERIENCE OF THE PCP INCLUDING DISCUSSING SPECIFIC PATIENT NAMES AND RESULTS]

1. Do you recall receiving the results for [PATIENT] via email? [Interviewer may show result report]
2. What was your initial reaction to receiving the result?
3. How helpful were the resources provided with the result [Interviewer may show treatment algorithm provided with results]
4. How prepared did you feel discussing these results with your patient(s)?
    a. Please describe what you reviewed with your patient
    b. Please describe what resources you used in your conversation with the patient.

5. What additional resources would have been helpful in discussing this result with your patient?
6. How would you describe the benefit of receiving genetic testing results via MVP-ROAR?
7. How would you describe the barriers to testing within MVP-ROAR?
8. How could the current return of results process be improved?



VA Central IRB
Effective Date: October 4, 2022

9. Would you like to see return of result efforts within MVP expanded to other conditions?
10. How would you prefer to receive these results in the future (email, fax, postal, CPRS alert etc.)

General [AGAIN, INTERVIEWER USES THESE PROMPTS AS A GENERAL GUIDE BUT MIGHT ASK RELEVANT FOLLOW-UP QUESTIONS AS APPROPRIATE]

1. What are the potential benefits of genetic testing?
    a. Does this differ when testing for at risk individuals (screening) vs. diagnostic?
2. What are potential barriers to offering genetic testing?
3. How do you think patients within VHA feel about receiving genetic testing results?
4. In general, how do you feel about return of results within MVP?
5. Describe your experience, if any, with familial hypercholesterolemia
6. How comfortable are you with managing FH? What resources/support would be helpful?

Do you have any additional comments you'd like to make about your experience with receiving genetic research results?

Thank you very much for your participation. That concludes the interview, and I will stop the recording.

[STOP RECORDING]



VA Central IRB
Effective Date: October 4, 2022
